# Single-Axis Fairness Interventions Produce Asymmetric Cross-Axis Effects in Clinical Prediction

**DOI:** 10.64898/2026.06.16.26355120

**Authors:** Keun Young Yoon, Hyunjung Gloria Kwak

## Abstract

**Objective:** To systematically evaluate whether single-axis demographic balancing introduces cross-axis fairness trade-offs in clinical prediction, and to characterize the directional asymmetry of these effects across model architectures and balancing strategies.

**Materials and Methods:** We evaluated cross-axis fairness effects of demographic balancing in one-year all-cause mortality prediction using the MIMIC-IV database (N=64,427). Seven machine learning architectures were trained under six balancing strategies targeting either gender or race, with performance assessed via outcome-stratified 80-20 train-test splits repeated 30 times across both targeted and non-targeted axes using AUC, TPR, and Brier score.

**Results:** Gender-targeting interventions largely preserved race fairness, while race-targeting consistently disrupted gender fairness across all methods and the majority of architectures. This asymmetry was invisible to same-axis evaluation alone. Race-targeting also incurred greater performance costs and calibration loss, with observed fairness gains potentially reflecting leveling down rather than genuine improvement. The same intervention could appear successful under TPR but fail under AUC evaluation.

**Discussion:** The asymmetry likely reflects differential category complexity: binary gender balancing requires modest distributional shifts, whereas multi-category race balancing necessitates aggressive reweighting that propagates to correlated axes. Cross-axis fairness effects are directionally dependent and metric-sensitive, indicating that single-metric, single-axis evaluation is insufficient.

**Conclusion:** Single-axis fairness optimization cannot guarantee cross-dimensional equity. Cross-axis, multi-metric fairness evaluation should be integrated into pre-deployment auditing of healthcare artificial intelligence (AI) models.

## BACKGROUND AND SIGNIFICANCE

Machine learning (ML) has become integral to modern healthcare; ^1,2^ yet these models often exhibit unequal performance across demographic groups, achieving high overall accuracy while producing systematically larger errors for certain subpopulations. ^3,4^ To address such disparities, researchers have proposed fairness interventions, such as class re-weighting and minority oversampling, to correct imbalances in training data. ^5,6^ However, these techniques are typically optimized and evaluated along a single demographic axis, ^7^ potentially introducing inequities at the intersections of demographic categories. ^8^ Consequently, an intervention that resolves disparities along one dimension may inadvertently leave unaddressed or even exacerbate disparities along others.

While prior work has documented algorithmic biases affecting underserved populations; ^9^ and theorized inherent tensions among fairness criteria, ^10^ most evaluations remain confined to a single demographic axis. ^11^ La Cava et al. ^12^ showed that marginal fairness gains do not reliably translate to intersectional subgroups, and Lett and La Cava ^7^ articulated intersectionality as a guiding framework for fair ML in health sciences. More recently, Chen et al. ^13^ demonstrated that single-attribute fairness methods degrade fairness on unconsidered attributes in up to 88% of scenarios in general ML benchmarks, and analogous effects have been observed in limited clinical contexts. ^14,15^ Yet no study has systematically evaluated how standard debiasing methods affect non-targeted axes across multiple model architectures and demographic dimensions in clinical prediction settings.

Given this gap, we investigate whether fairness interventions designed along a single demographic axis can achieve equity across multiple dimensions simultaneously in clinical prediction. Specifically, we ask: (1) Does single-axis fairness balancing improve equity on the targeted demographic dimension without degrading predictive performance? (2) Do improvements on one axis inadvertently worsen fairness along non-targeted axes (cross-axis trade-offs)? (3) How do intersectional subgroups fare under different balancing strategies?

To answer these questions, we evaluated seven ML architectures - spanning both traditional ML, gradient boosting, and deep tabular models - and trained under six fairness-oriented balancing strategies on one-year all-cause mortality prediction using the Medical Information Mart for Intensive Care IV (MIMIC-IV) database. Each strategy targeted a single demographic axis (gender or race), and model performance was assessed in terms of discrimination (Area Under the Receiver Operating Characteristic Curve [AUC]), calibration (Brier score), and fairness (True Positive Rate [TPR], False Positive Rate [FPR], and Positive Prediction Rate [PPR] gaps) across both the targeted and non-targeted axes. Our results reveal a consistent asymmetry: interventions targeting binary categories (gender) maintain neutrality on multi-category axes (race), whereas the reverse does not hold.

This study provides (1) a systematic cross-axis evaluation of fairness interventions across model families, (2) empirical evidence that single-axis mitigation can reduce disparities locally yet degrade fairness along non-targeted axes, and (3) a cross-axis evaluation framework for multidimensional fairness assessment in healthcare AI models.

## METHODS

### Dataset and Study Population

We used the MIMIC-IV version 3.1 database, ^16^ a publicly available dataset containing de-identified electronic health records from patients admitted to the Beth Israel Deaconess Medical Center, Boston, MA, USA, from 2008 to 2022. This study used de-identified data under PhysioNet Credentialed Access and was exempt from institutional review board approval.

Eligible patients were adults (age ≥18 years) with at least one intensive care unit (ICU) admission who met all of the following conditions: (1) minimum hospital length of stay of 24 hours, (2) survival beyond the first 24 hours of hospital admission to minimize bias from immediately fatal cases, and (3) at least one ICU stay during the hospitalization.

For patients with multiple qualifying admissions, we selected the most recent admission with an ICU stay as the index admission to avoid within-patient correlation. Within each index admission, the first ICU stay was used as the analytic unit. A total of 66,056 unique patients met these inclusion criteria.

We applied the following exclusion criteria: (1) more than 50% of core clinical variables missing (hemoglobin, white blood cell count, creatinine, Charlson Comorbidity Index [CCI], age; n=1,428), and (2) physiologically implausible vital sign values (peripheral oxygen saturation [SpO_2_] >100%, respiratory rate >60/min, systolic blood pressure >300 or <30 mmHg, heart rate >300/min; n=207). After exclusions (n=1,629 unique patients; 6 patients met both criteria), the final analytic cohort comprised 64,427 patients with 22,260 (34.6%) mortality events.

Demographic features included sex, race, age at admission, admission type (elective vs. emergency), and insurance status. Clinical variables including vital signs (heart rate, blood pressure, respiratory rate, temperature, oxygen saturation, and Glasgow Coma Scale [GCS]), which overlap substantially with features used in established ICU prediction benchmarks, ^17^ and laboratory values (creatinine, hemoglobin, white blood cell count, blood glucose, and lactate) consistent with variables in validated ICU severity scoring systems, ^18^ were extracted from the first 24 hours following the first ICU stay of the index admission to capture early clinical presentation and ensure standardized data availability across patients. Race/ethnicity was categorized into eight categories: White, Black, Asian, Hispanic, Pacific Islander, Native American, Other (including patients with multiple race/ethnicity designations), and Unknown (including missing race data). Classification was determined using the most recent admission with a valid (non-missing, non-unknown) race designation for each patient.

### Outcome Definition

The primary outcome was one-year all-cause mortality, defined as death within 365 days of hospital admission. We selected this over in-hospital or ICU mortality to ensure sufficient event counts across demographic subgroups for reliable fairness estimation; the rationale is detailed in Supplementary Methods. Sensitivity analyses using in-hospital and ICU mortality confirmed similar cross-axis patterns (Table S2).

### Data Preprocessing

Patients without a recorded death date were classified as survivors. Binary missingness indicators were generated for all vital sign, laboratory, and GCS variables prior to imputation and included as additional features. Remaining missing values were imputed using median (continuous) or mode (categorical) imputation. Continuous features were z-score normalized across all models. High-cardinality categorical variables were reduced to top categories plus “Other” and one-hot encoded.

### Models

We employed **seven** predictive architectures spanning traditional machine learning, gradient boosting, and deep learning approaches: Logistic Regression (LR), Extreme Gradient Boosting (XG-Boost), ^19^ Multilayer Perceptron (MLP), TabNet, ^20^ FT-Transformer, ^21^ TabPFN, ^22^ and Explainable Boosting Machine (EBM). ^23^ This diversity enables assessment of whether fairness intervention effects generalize across model architectures. Hyperparameter configurations are provided in Table S1.

#### Balancing Strategies

We compared each baseline model with **six** balancing strategies applied along a single demographic axis (gender or race):

- **Baseline**: No modification to the original data distribution.
- **Resampling Methods**: Oversampling (OS) duplicates underrepresented subgroup instances with replacement; undersampling (US) removes majority subgroup instances to match the smallest group size.
- **Class Weighting (CW)**: Instances weighted inversely proportional to subgroup frequency: *w*_*g*_ = *N*/(*K* × *n*_*g*_).
- **Group DRO (GD)** ^24^: Robust optimization minimizing worst-case loss across subgroups via dynamic reweighting during training.
- **Adversarial Debiasing (ADV)**^25^: In-processing method using an adversary network to penalize demographic information leakage from predictions.
- **Adaptive Balanced Sampling (ADB)**: Joint weighting scheme that assigns instance weights inversely proportional to the frequency of each demographic-outcome combination: *w*_*g,y*_ = 1/(2 × *K* × *n*_*g,y*_), where *K* is the number of demographic groups and *n*_*g,y*_ is the count of samples in group *g* with outcome *y*.

#### Experimental Protocol

For each balancing strategy, we conducted separate experiments balancing by gender and by race to evaluate cross-dimensional fairness effects. All models were trained and evaluated under outcome-stratified 80–20 train-test splits with 30 random repetitions for statistical robustness. Due to computational constraints, EBM training was limited to 15,000 randomly sampled instances (approximately 29% of the training set), and TabNet to 50,000 (approximately 97%); all other models used the full training set. We report mean performance and 95% confidence intervals (CIs) across splits. 95% confidence intervals are provided for overall AUC (Table 2); all other tables report mean values.

### Statistical Analysis

Continuous variables were described as mean (SD) or median [IQR] after normality assessment using the Shapiro-Wilk test. Normally distributed continuous variables were compared between survivors and non-survivors using Student’s *t*-test; non-normally distributed variables using the Mann-Whitney U test (two-sided). Categorical variables were compared using the chi-square test. All analyses were performed using Python 3.9.0 with SciPy 1.13.1 and scikit-learn 1.6.1.

### Evaluation Framework

We systematically evaluated model performance and fairness at three levels: (1) targeted axes, subgroups within the balanced dimension (e.g., gender subgroups when balancing by gender), (2) non-targeted axes, subgroups in dimensions not subject to balancing (e.g., racial subgroups when balancing by gender), and (3) intersectional subgroups defined by gender–race combinations (e.g., Gender_*g*_ × Race_*r*_).

For each demographic subgroup *g*, we computed AUC as the primary performance metric, with calibration assessed via Brier Score. For fairness evaluation, we computed TPR, FPR, and PPR at the default classification threshold of 0.5. To quantify disparities across subgroups, we computed gap measures as the maximum pairwise difference across subgroups within a given demographic axis: ΔAUC, ΔTPR, and analogously for ΔFPR and ΔPPR.

For race, gaps were computed over six primary groups (*S*_race_ = {White, Black, Asian, Hispanic, Other, Unknown}); Native American (n=127) and Pacific Islander (n=99) were excluded owing to small sample sizes (n < 130; 95% CI width > 0.20), as reliable fairness metric estimation requires sufficient subgroup representation. ^26^

#### Single-Axis Evaluation

We first assessed whether each balancing strategy reduced ΔAUC and ΔTPR on the targeted demographic axis relative to the unbalanced baseline.

#### Cross-Axis Evaluation

Our cross-axis evaluation examines how balancing along one demographic dimension affects fairness along non-targeted dimensions: when balancing by gender, we evaluated fairness across racial subgroups, and conversely, when balancing by race, we assessed fairness across gender subgroups. To classify the joint outcome of each model-method combination across both axes, we defined four categories based on the direction of change in ΔAUC and ΔTPR relative to baseline: Win-win (W): both targeted and non-targeted axes improved; Trade-off (T): targeted axis improved, non-targeted axis worsened; Paradox (P): targeted axis worsened, non-targeted axis improved; Both-worse (B): both axes worsened.

#### Intersectional Evaluation

Intersectional subgroup performance was evaluated separately using AUC across gender-race combinations (Gender_*g*_ × Race_*r*_). Intersectional subgroups serve as an evaluation lens rather than optimization targets; all fairness interventions operate along a single demographic axis at a time.

## RESULTS

We evaluated seven machine learning architectures across six demographic balancing methods with 30 random train-test splits on MIMIC-IV critical care data (N=64,427; one-year mortality rate=34.6%). This section presents our findings organized around three central research questions: whether single-axis balancing achieves intended fairness improvements, whether cross-axis trade-offs emerge, and how intersectional subgroups are affected.

### Cohort Characteristics

After applying inclusion and exclusion criteria, our final analytic cohort comprised 64,427 unique ICU admissions, with 22,260 (34.6%) mortality events. The mean age was 65.4 years (SD 16.8), and 43.3% were Female.

The racial distribution exhibited substantial imbalance: White patients comprised 67.4% of the cohort, followed by Black (8.9%), Hispanic (3.6%), and Asian (3.0%), while Pacific Islander (0.2%) and Native American (0.2%) groups were notably underrepresented. When examining intersectional subgroups defined by gender and race, this imbalance was further amplified; the largest group (Male White, n=24,720) outnumbered the smallest group (Female Pacific Islande, n=33) by approximately 749-fold (Figure 1).

**Figure 1.**
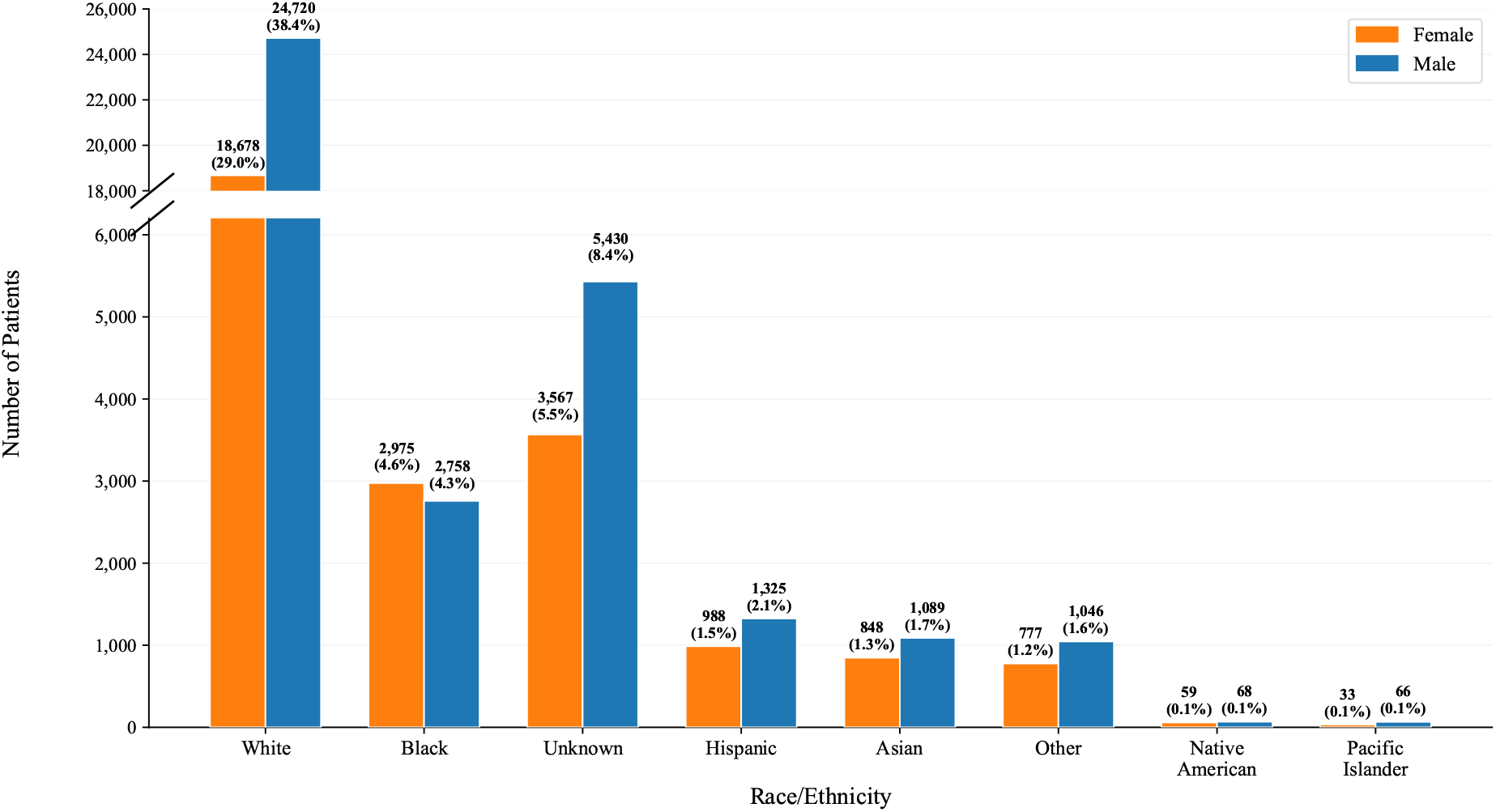
Distribution of the study population by gender and race/ethnicity (N = 64,427). The y-axis is broken to accommodate the extreme imbalance between the majority group (White in the figure) and all other groups. White patients comprised the largest group (67.4%), followed by Unknown (14.0%), Black (8.9%), Hispanic (3.6%), Asian (3.0%), Other (2.8%), Native American (0.2%), and Pacific Islander (0.2%). Overall, 56.7% of patients were Male (n = 36,502) and 43.3% were Female (n = 27,925). Male predominance was observed across most racial/ethnic categories, with the exception of Black patients (F = 2,975 vs. M = 2,758). *Alt text: Grouped bar chart with a broken y-axis showing patient counts by race and gender. White patients dominate with 18,678 females and 24,720 males. Unknown is the next largest group with 3,567 females and 5,430 males. Black follows with 2,975 females and 2,758 males, the only category in which females outnumber males. All remaining groups have fewer than 2,500 patients each, and Native American and Pacific Islander groups have fewer than 70 patients per gender*.

Mortality was associated with older age (71.8 vs. 62.0 years), higher comorbidity burden (CCI 7.6 vs. 4.6), longer ICU duration (median 70.2 vs. 46.3 hours), and Medicare insurance (69.9% vs. 47.0%). The mortality rate was higher among Female (36.0%) than Male patients (33.5%), indicating that demographic imbalances extend beyond group size to outcome prevalence. Overall clinical and demographic characteristics are summarized in Table 1.

**Table 1.** Baseline Characteristics of the Study Cohort Stratified by Mortality Status.

| Variable | Missing (%) | Overall (n=64,427) | Survival (n=42,167) | Death (n=22,260) | p-value |
| --- | --- | --- | --- | --- | --- |
| Age (years), mean (SD) | 0.0 | 65.4 (16.8) | 62.0 (17.0) | 71.8 (14.3) | <0.001 |
| **Gender, n (%)** | 0.0 |  |  |  | <0.001 |
| Male |  | 36,502 (56.7) | 24,289 (57.6) | 12,213 (54.9) |  |
| Female |  | 27,925 (43.3) | 17,878 (42.4) | 10,047 (45.1) |  |
| **Race, n (%)** | 0.0 |  |  |  | <0.001 |
| White |  | 43,398 (67.4) | 28,059 (66.5) | 15,339 (68.9) |  |
| Black |  | 5,733 (8.9) | 3,468 (8.2) | 2,265 (10.2) |  |
| Hispanic |  | 2,313 (3.6) | 1,700 (4.0) | 613 (2.8) |  |
| Asian |  | 1,937 (3.0) | 1,273 (3.0) | 664 (3.0) |  |
| Other |  | 1,823 (2.8) | 1,354 (3.2) | 469 (2.1) |  |
| Native American |  | 127 (0.2) | 94 (0.2) | 33 (0.1) |  |
| Pacific Islander |  | 99 (0.2) | 67 (0.2) | 32 (0.1) |  |
| Unknown |  | 8,997 (14.0) | 6,152 (14.6) | 2,845 (12.8) |  |
| **Insurance, n (%)** | 1.7 |  |  |  | <0.001 |
| Medicare |  | 35,384 (54.9) | 19,825 (47.0) | 15,559 (69.9) |  |
| Private |  | 17,216 (26.7) | 13,782 (32.7) | 3,434 (15.4) |  |
| Medicaid |  | 9,135 (14.2) | 6,424 (15.2) | 2,711 (12.2) |  |
| Other |  | 1,622 (2.5) | 1,293 (3.1) | 329 (1.5) |  |
| Charlson Comorbidity Index, mean (SD) | 0.0 | 5.6 (3.1) | 4.6 (2.7) | 7.6 (2.9) | <0.001 |
| GCS Total (min), median [IQR] | 0.0 | 15.0 [14.0, 15.0] | 15.0 [14.0, 15.0] | 15.0 [13.0, 15.0] | <0.001 |
| Length of Stay (hrs), median [IQR] | 0.0 | 183.0 [108.0, 333.0] | 164.0 [102.0, 282.0] | 233.0 [122.0, 440.0] | <0.001 |
| ICU Duration (hrs), median [IQR] | 0.0 | 51.5 [29.0, 104.0] | 46.3 [27.3, 85.5] | 70.2 [36.7, 150.6] | <0.001 |

*Note:* Continuous variables reported as mean (SD) or median [IQR] based on distribution; normality assessed using the Shapiro-Wilk test; normally distributed variables compared using Student's *t*-test, non-normally distributed variables using the Mann-Whitney U test (two-sided). Categorical variables compared using the chi-square test. GCS, Glasgow Coma Scale; ICU, Intensive Care Unit; IQR, Interquartile Range; SD, Standard Deviation.

**Table 2.**
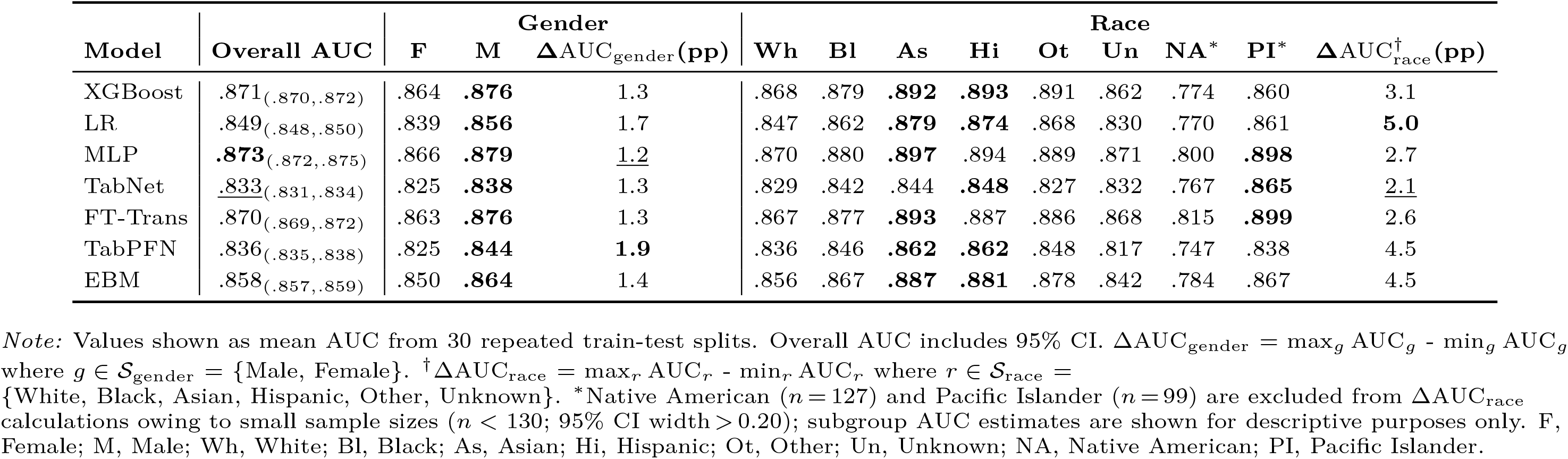
Baseline Model Performance and Pre-Existing Demographic Disparities for One-Year All-Cause Mortality Prediction.

| Model | Overall AUC | Gender | | | Race | | | | | | | | $\Delta AUC_{\text{race}}^{\dagger}(\text{pp})$ |
| --- | --- | --- | --- | --- | --- | --- | --- | --- | --- | --- | --- | --- | --- |
| | | F | M | $\Delta AUC_{\text{gender}}(\text{pp})$ | Wh | Bl | As | Hi | Ot | Un | NA* | PI* | |
| XGBoost | .871(.870,.872) | .864 | **.876** | 1.3 | .868 | .879 | **.892** | **.893** | .891 | .862 | .774 | .860 | 3.1 |
| LR | .849(.848,.850) | .839 | **.856** | 1.7 | .847 | .862 | **.879** | **.874** | .868 | .830 | .770 | .861 | **5.0** |
| MLP | **.873**(.872,.875) | .866 | **.879** | <u>1.2</u> | .870 | .880 | **.897** | .894 | .889 | .871 | .800 | **.898** | 2.7 |
| TabNet | <u>.833</u>(.831,.834) | .825 | **.838** | 1.3 | .829 | .842 | .844 | **.848** | .827 | .832 | .767 | **.865** | <u>2.1</u> |
| FT-Trans | .870(.869,.872) | .863 | **.876** | 1.3 | .867 | .877 | **.893** | .887 | .886 | .868 | .815 | **.899** | 2.6 |
| TabPFN | .836(.835,.838) | .825 | **.844** | **1.9** | .836 | .846 | **.862** | **.862** | .848 | .817 | .747 | .838 | 4.5 |
| EBM | .858(.857,.859) | .850 | **.864** | 1.4 | .856 | .867 | **.887** | **.881** | .878 | .842 | .784 | .867 | 4.5 |

Note: Values shown as mean AUC from 30 repeated train-test splits. Overall AUC includes 95% CI. $\Delta AUC_{\text{gender}} = \max_g AUC_g - \min_g AUC_g$ where $g \in \mathcal{S}_{\text{gender}} = \{\text{Male, Female}\}$ . $\dagger \Delta AUC_{\text{race}} = \max_r AUC_r - \min_r AUC_r$ where $r \in \mathcal{S}_{\text{race}} = \{\text{White, Black, Asian, Hispanic, Other, Unknown}\}$ . \*Native American ( $n=127$ ) and Pacific Islander ( $n=99$ ) are excluded from $\Delta AUC_{\text{race}}$ calculations owing to small sample sizes ( $n < 130$ ; 95% CI width $> 0.20$ ); subgroup AUC estimates are shown for descriptive purposes only. F, Female; M, Male; Wh, White; Bl, Black; As, Asian; Hi, Hispanic; Ot, Other; Un, Unknown; NA, Native American; PI, Pacific Islander.

### Baseline Model Performance

Prior to any fairness intervention, we established baseline predictive performance and pre-existing demographic disparities across all seven models (Table 2). Overall AUC ranged from 0.833 [0.831–0.834] for Tab-Net to 0.873 [0.872–0.875] for MLP. Narrow CIs across splits indicate stable performance rather than sampling variability.

All seven models exhibited pre-existing gender disparities, with ΔAUC_gender_ ranging from 1.2 percentage points (pp) in MLP to 1.9 pp in TabPFN. Male AUC exceeded Female AUC across all architectures.

Pre-existing race disparities were greater: ΔAUC_race_ ranged from 2.1 pp in TabNet to 5.0 pp in LR. Subgroup-level AUC followed consistent pairwise patterns across all seven models: Asian and Hispanic patients showed the highest AUC (except NA and PI), Black exceeded White, and Other exceeded Unknown in six of seven models. The relative ranking of Asian and Hispanic varied by architecture. Notably, ΔAUC_race_ consistently exceeded ΔAUC_gender_, foreshadowing the greater distributional shifts required by race-targeting interventions.

### Single-Axis Effects

We first examined whether demographic balancing methods achieved their intended effect of reducing fairness disparities on the targeted axis. Following standard fairness evaluation practices, we assessed both AUC parity (equal predictive performance) and TPR parity (equal sensitivity) across demographic groups.

#### Gender-Targeting

Baseline gender fairness gaps were small (ΔAUC_gender_: 1.2–1.9 pp, Table 3 Panels A; ΔTPR_gender_: 0.004–0.027, Table 4 Panels A). Simple resampling methods (OS, US, CW) narrowed ΔAUC_gender_ in 5–7 of 7 models but ΔTPR_gender_ in at most 2 of 7. Advanced methods (GD, ADV, and ADB) narrowed ΔTPR_gender_ in 3–4 of 7 models but ΔAUC_gender_ in 2–4 of 7.

**Table 3.**
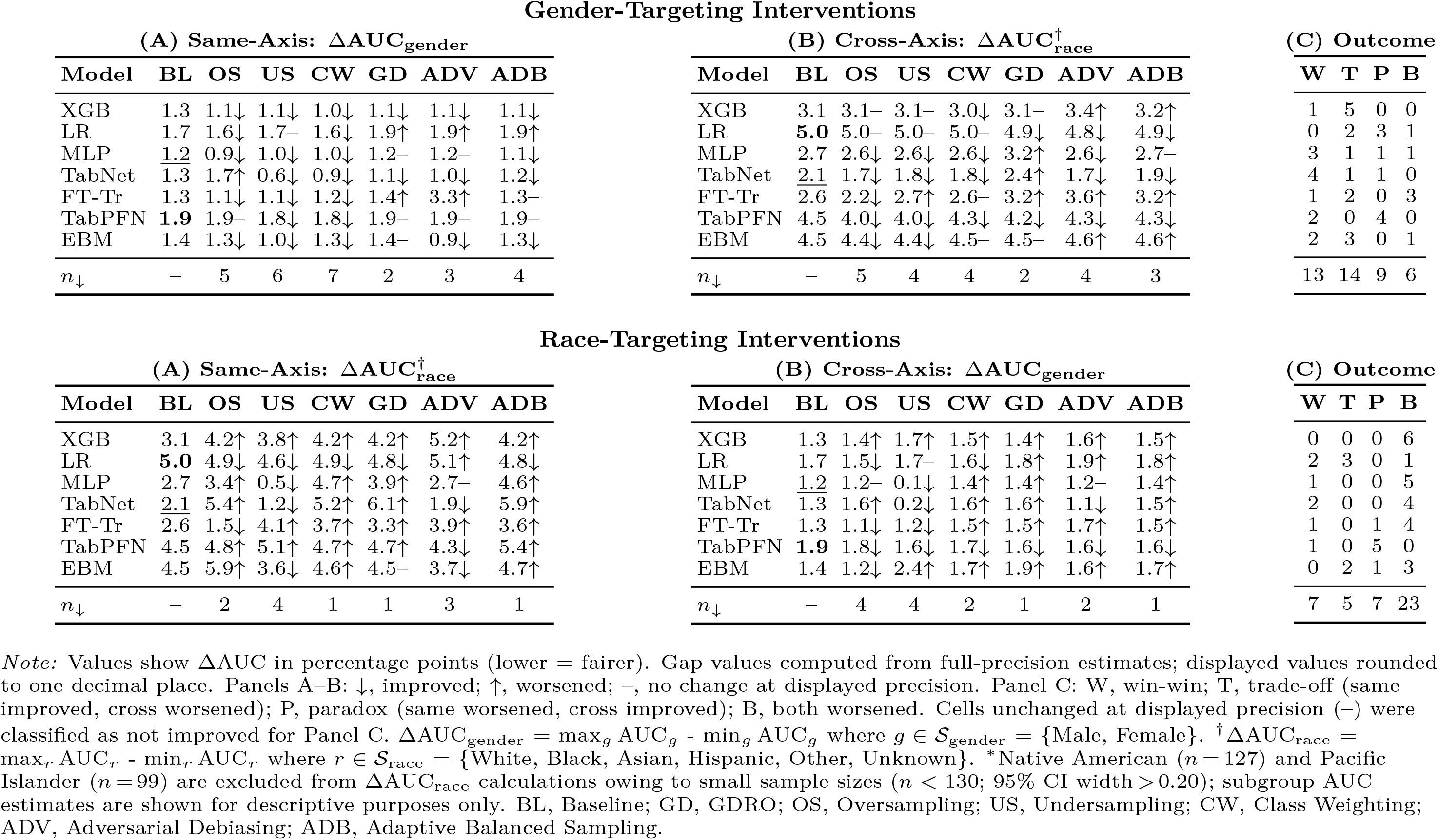
Effects of Single-Axis Targeting Interventions on Same-Axis and Cross-Axis AUC Gaps. Panel (A), same-axis; Panel (B), cross-axis; Panel (C), joint outcome classification.

| Gender-Targeting Interventions |  |  |  |  |  |  |  |  |  |  |  |  |  |  |  |  |  |  |  |
| --- | --- | --- | --- | --- | --- | --- | --- | --- | --- | --- | --- | --- | --- | --- | --- | --- | --- | --- | --- |
| (A) Same-Axis: $\Delta\text{AUC}_{\text{gender}}$ | | | | | | | | (B) Cross-Axis: $\Delta\text{AUC}_{\text{race}}^{\dagger}$ | | | | | | | | (C) Outcome | | | |
| Model | BL | OS | US | CW | GD | ADV | ADB | Model | BL | OS | US | CW | GD | ADV | ADB | W | T | P | B |
| XGB | 1.3 | 1.1↓ | 1.1↓ | 1.0↓ | 1.1↓ | 1.1↓ | 1.1↓ | XGB | 3.1 | 3.1- | 3.1- | 3.0↓ | 3.1- | 3.4↑ | 3.2↑ | 1 | 5 | 0 | 0 |
| LR | 1.7 | 1.6↓ | 1.7- | 1.6↓ | 1.9↑ | 1.9↑ | 1.9↑ | LR | **5.0** | 5.0- | 5.0- | 5.0- | 4.9↓ | 4.8↓ | 4.9↓ | 0 | 2 | 3 | 1 |
| MLP | 1.2 | 0.9↓ | 1.0↓ | 1.0↓ | 1.2- | 1.2- | 1.1↓ | MLP | 2.7 | 2.6↓ | 2.6↓ | 2.6↓ | 3.2↑ | 2.6↓ | 2.7- | 3 | 1 | 1 | 1 |
| TabNet | 1.3 | 1.7↑ | 0.6↓ | 0.9↓ | 1.1↓ | 1.0↓ | 1.2↓ | TabNet | 2.1 | 1.7↓ | 1.8↓ | 1.8↓ | 2.4↑ | 1.7↓ | 1.9↓ | 4 | 1 | 1 | 0 |
| FT-Tr | 1.3 | 1.1↓ | 1.1↓ | 1.2↓ | 1.4↑ | 3.3↑ | 1.3- | FT-Tr | 2.6 | 2.2↓ | 2.7↑ | 2.6- | 3.2↑ | 3.6↑ | 3.2↑ | 1 | 2 | 0 | 3 |
| TabPFN | **1.9** | 1.9- | 1.8↓ | 1.8↓ | 1.9- | 1.9- | 1.9- | TabPFN | 4.5 | 4.0↓ | 4.0↓ | 4.3↓ | 4.2↓ | 4.3↓ | 4.3↓ | 2 | 0 | 4 | 0 |
| EBM | 1.4 | 1.3↓ | 1.0↓ | 1.3↓ | 1.4- | 0.9↓ | 1.3↓ | EBM | 4.5 | 4.4↓ | 4.4↓ | 4.5- | 4.5- | 4.6↑ | 4.6↑ | 2 | 3 | 0 | 1 |
| $n_{\downarrow}$ | - | 5 | 6 | 7 | 2 | 3 | 4 | $n_{\downarrow}$ | - | 5 | 4 | 4 | 2 | 4 | 3 | 13 | 14 | 9 | 6 |

| Race-Targeting Interventions |  |  |  |  |  |  |  |  |  |  |  |  |  |  |  |  |  |  |  |
| --- | --- | --- | --- | --- | --- | --- | --- | --- | --- | --- | --- | --- | --- | --- | --- | --- | --- | --- | --- |
| (A) Same-Axis: $\Delta\text{AUC}_{\text{race}}^{\dagger}$ | | | | | | | | (B) Cross-Axis: $\Delta\text{AUC}_{\text{gender}}$ | | | | | | | | (C) Outcome | | | |
| Model | BL | OS | US | CW | GD | ADV | ADB | Model | BL | OS | US | CW | GD | ADV | ADB | W | T | P | B |
| XGB | 3.1 | 4.2↑ | 3.8↑ | 4.2↑ | 4.2↑ | 5.2↑ | 4.2↑ | XGB | 1.3 | 1.4↑ | 1.7↑ | 1.5↑ | 1.4↑ | 1.6↑ | 1.5↑ | 0 | 0 | 0 | 6 |
| LR | **5.0** | 4.9↓ | 4.6↓ | 4.9↓ | 4.8↓ | 5.1↑ | 4.8↓ | LR | 1.7 | 1.5↓ | 1.7- | 1.6↓ | 1.8↑ | 1.9↑ | 1.8↑ | 2 | 3 | 0 | 1 |
| MLP | 2.7 | 3.4↑ | 0.5↓ | 4.7↑ | 3.9↑ | 2.7- | 4.6↑ | MLP | 1.2 | 1.2- | 0.1↓ | 1.4↑ | 1.4↑ | 1.2- | 1.4↑ | 1 | 0 | 0 | 5 |
| TabNet | 2.1 | 5.4↑ | 1.2↓ | 5.2↑ | 6.1↑ | 1.9↓ | 5.9↑ | TabNet | 1.3 | 1.6↑ | 0.2↓ | 1.6↑ | 1.6↑ | 1.1↓ | 1.5↑ | 2 | 0 | 0 | 4 |
| FT-Tr | 2.6 | 1.5↓ | 4.1↑ | 3.7↑ | 3.3↑ | 3.9↑ | 3.6↑ | FT-Tr | 1.3 | 1.1↓ | 1.2↓ | 1.5↑ | 1.5↑ | 1.7↑ | 1.5↑ | 1 | 0 | 1 | 4 |
| TabPFN | 4.5 | 4.8↑ | 5.1↑ | 4.7↑ | 4.7↑ | 4.3↓ | 5.4↑ | TabPFN | **1.9** | 1.8↓ | 1.6↓ | 1.7↓ | 1.6↓ | 1.6↓ | 1.6↓ | 1 | 0 | 5 | 0 |
| EBM | 4.5 | 5.9↑ | 3.6↓ | 4.6↑ | 4.5- | 3.7↓ | 4.7↑ | EBM | 1.4 | 1.2↓ | 2.4↑ | 1.7↑ | 1.9↑ | 1.6↑ | 1.7↑ | 0 | 2 | 1 | 3 |
| $n_{\downarrow}$ | - | 2 | 4 | 1 | 1 | 3 | 1 | $n_{\downarrow}$ | - | 4 | 4 | 2 | 1 | 2 | 1 | 7 | 5 | 7 | 23 |

Note: Values show $\Delta\text{AUC}$ in percentage points (lower = fairer). Gap values computed from full-precision estimates; displayed values rounded to one decimal place. Panels A-B: ↓, improved; ↑, worsened; -, no change at displayed precision. Panel C: W, win-win; T, trade-off (same improved, cross worsened); P, paradox (same worsened, cross improved); B, both worsened. Cells unchanged at displayed precision (-) were classified as not improved for Panel C. $\Delta\text{AUC}_{\text{gender}} = \max_g \text{AUC}_g - \min_g \text{AUC}_g$ where $g \in S_{\text{gender}} = \{\text{Male, Female}\}$ . $\dagger\Delta\text{AUC}_{\text{race}} = \max_r \text{AUC}_r - \min_r \text{AUC}_r$ where $r \in S_{\text{race}} = \{\text{White, Black, Asian, Hispanic, Other, Unknown}\}$ . \*Native American ( $n=127$ ) and Pacific Islander ( $n=99$ ) are excluded from $\Delta\text{AUC}_{\text{race}}$ calculations owing to small sample sizes ( $n < 130$ ; 95% CI width $> 0.20$ ); subgroup AUC estimates are shown for descriptive purposes only. BL, Baseline; GD, GDRO; OS, Oversampling; US, Undersampling; CW, Class Weighting; ADV, Adversarial Debiasing; ADB, Adaptive Balanced Sampling.

**Table 4.** Effects of Single-Axis Targeting Interventions on Same-Axis and Cross-Axis TPR Gaps. Panel (A), same-axis; Panel (B), cross-axis; Panel (C), joint outcome classification.

| Gender-Targeting Interventions |  |  |  |  |  |  |  |  |  |  |  |  |  |  |  |  |  |  |  |
| --- | --- | --- | --- | --- | --- | --- | --- | --- | --- | --- | --- | --- | --- | --- | --- | --- | --- | --- | --- |
| (A) Same-Axis: $\Delta\text{TPR}_{\text{gender}}$ | | | | | | | | (B) Cross-Axis: $\Delta\text{TPR}_{\text{race}}^{\dagger}$ | | | | | | | | (C) Outcome | | | |
| Model | BL | OS | US | CW | GD | ADV | ADB | Model | BL | OS | US | CW | GD | ADV | ADB | W | T | P | B |
| XGB | **.027** | .028↑ | .027- | .029↑ | .009↓ | .021↓ | .011↓ | XGB | .141 | .143↑ | .142↑ | .144↑ | .089↓ | .137↓ | .095↓ | 3 | 0 | 0 | 3 |
| LR | .008 | .008- | .007↓ | .008- | .024↑ | .039↑ | .023↑ | LR | .170 | .170- | .175↑ | .176↑ | .131↓ | .158↓ | .130↓ | 0 | 1 | 3 | 2 |
| MLP | .013 | .019↑ | .014↑ | .019↑ | .007↓ | .005↓ | .002↓ | MLP | .132 | .120↓ | .130↓ | .127↓ | .099↓ | .140↑ | .100↓ | 2 | 1 | 3 | 0 |
| TabNet | .007 | .011↑ | .010↑ | .003↓ | .011↑ | .005↓ | .014↑ | TabNet | <u>.104</u> | .095↓ | .094↓ | .097↓ | .099↓ | .089↓ | .094↓ | 2 | 0 | 4 | 0 |
| FT-Tr | .005 | .018↑ | .008↑ | .013↑ | .014↑ | .020↑ | .012↑ | FT-Tr | .126 | .120↓ | .137↑ | .128↑ | .120↓ | .135↑ | .119↓ | 0 | 0 | 3 | 3 |
| TabPFN | .010 | .012↑ | .001↓ | .007↓ | .006↓ | .004↓ | .008↓ | TabPFN | **.230** | .216↓ | .206↓ | .188↓ | .152↓ | .169↓ | .171↓ | 5 | 0 | 1 | 0 |
| EBM | <u>.004</u> | .004- | .004- | .005↑ | .005↑ | .011↑ | .005↑ | EBM | .181 | .171↓ | .167↓ | .184↑ | .145↓ | .181- | .140↓ | 0 | 0 | 4 | 2 |
| $n_{\downarrow}$ | - | 0 | 2 | 2 | 3 | 4 | 3 | $n_{\downarrow}$ | - | 5 | 4 | 3 | 7 | 4 | 7 | 12 | 2 | 18 | 10 |

| Race-Targeting Interventions |  |  |  |  |  |  |  |  |  |  |  |  |  |  |  |  |  |  |  |
| --- | --- | --- | --- | --- | --- | --- | --- | --- | --- | --- | --- | --- | --- | --- | --- | --- | --- | --- | --- |
| (A) Same-Axis: $\Delta\text{TPR}_{\text{race}}^{\dagger}$ | | | | | | | | (B) Cross-Axis: $\Delta\text{TPR}_{\text{gender}}$ | | | | | | | | (C) Outcome | | | |
| Model | BL | OS | US | CW | GD | ADV | ADB | Model | BL | OS | US | CW | GD | ADV | ADB | W | T | P | B |
| XGB | .141 | .150↑ | .133↓ | .149↑ | .076↓ | .173↑ | .075↓ | XGB | **.027** | .021↓ | .003↓ | .021↓ | .022↓ | .018↓ | .022↓ | 3 | 0 | 3 | 0 |
| LR | .170 | .160↓ | .149↓ | .162↓ | .047↓ | .121↓ | .047↓ | LR | .008 | .051↑ | .038↑ | .052↑ | .035↑ | .015↑ | .035↑ | 0 | 6 | 0 | 0 |
| MLP | .132 | .105↓ | .014↓ | .143↑ | .017↓ | .035↓ | .019↓ | MLP | .013 | .001↓ | .008↓ | .009↓ | .009↓ | .011↓ | .002↓ | 5 | 0 | 1 | 0 |
| TabNet | <u>.104</u> | .100↓ | .023↓ | .093↓ | .094↓ | .068↓ | .088↓ | TabNet | .007 | .004↓ | .004↓ | .001↓ | .001↓ | .003↓ | .005↓ | 6 | 0 | 0 | 0 |
| FT-Tr | .126 | .080↓ | .114↓ | .142↑ | .030↓ | .058↓ | .030↓ | FT-Tr | .005 | .005- | .003↓ | .005- | .010↑ | .000↓ | .013↑ | 2 | 3 | 0 | 1 |
| TabPFN | **.230** | .168↓ | .169↓ | .157↓ | .101↓ | .158↓ | .100↓ | TabPFN | .010 | .032↑ | .059↑ | .029↑ | .020↑ | .016↑ | .016↑ | 0 | 6 | 0 | 0 |
| EBM | .181 | .135↓ | .202↑ | .163↓ | .056↓ | .177↓ | .052↓ | EBM | <u>.004</u> | .012↑ | .053↑ | .031↑ | .005↑ | .029↑ | .013↑ | 0 | 5 | 0 | 1 |
| $n_{\downarrow}$ | - | 6 | 6 | 4 | 7 | 6 | 7 | $n_{\downarrow}$ | - | 3 | 4 | 3 | 3 | 4 | 3 | 16 | 20 | 4 | 2 |

Note: Values show $\Delta\text{TPR}$ (lower = fairer). $\Delta\text{TPR}_{\text{gender}} = |\text{TPR}_{g1} - \text{TPR}_{g2}|$ where $g \in S_{\text{gender}} = \{\text{Male, Female}\}$ ; $\dagger\Delta\text{TPR}_{\text{race}} = \max_{r \in S_{\text{race}}} \text{TPR}_r - \min_{r \in S_{\text{race}}} \text{TPR}_r$ where $S_{\text{race}} = \{\text{White, Black, Asian, Hispanic, Other, Unknown}\}$ . Direction symbols, definitions, and abbreviations as in Table 3. Direction classifications reflect mean differences across 30 repeated splits; the majority of individual cell-level changes fall within sampling variability (95% CI). Aggregate pattern-level conclusions are supported by robust cases exceeding $1.96 \times \text{SE}$ : all robust gender-targeting cross-axis changes were improvements, while the majority of robust race-targeting cross-axis changes were worsening.

#### Race-Targeting

Baseline race fairness gaps were substantially larger (ΔAUC_race_: 2.1–5.0 pp, Table 3 Panels A; ΔTPR_race_: 0.104–0.230, Table 4 Panels A). Simple resampling methods (OS, US, CW) decreased ΔTPR_race_ in 4–6 of 7 models but ΔAUC_race_ in 1–4 of 7. Advanced methods decreased ΔTPR_race_ more consistently (GD and ADB: 7 each; ADV: 6) but widened ΔAUC_race_ in the majority, most consistently under GD (5 of 7) and ADB (6 of 7). These patterns indicate that same-axis fairness conclusions depend on the metric chosen, a divergence that also manifests in cross-axis evaluations (Section Cross-Axis Trade-offs).

Race-targeting also incurred greater overall performance costs than gender-targeting, including model collapse under US in two architectures (Table S3).

### Cross-Axis Trade-offs

Our central finding concerns the asymmetric cross-axis effects of single-axis demographic balancing (Table 3 and Table 4, Panels B and C).

#### Gender-Targeting

Gender-targeting interventions exhibited metric-dependent cross-axis effects on race fairness. Under AUC evaluation (Table 3, Panel B), simple methods (OS, US, CW) worsened (increased) ΔAUC_race_ in 0–1 of 7, while advanced methods worsened 3 of 7. Under TPR evaluation (Table 4, Panel B), the pattern reversed: simple resampling methods worsened ΔTPR_race_ in 1–4 of 7 models, while advanced methods worsened 0–2 of 7.

Panel C classifications in Table 3 and Table 4 confirmed this pattern. Under AUC, trade-off was the most frequent outcome (T=14 of 42 combinations [7 models × 6 methods]), with win-win (W=13), paradox (P=9), and both-worsened (B=6). Under TPR (Table 4), the dominant outcome was Paradox (P=18 of 42), reflecting that same-axis gender fairness rarely improved, yet cross-axis race fairness was also rarely harmed. Win-win (W) outcomes occurred in 12, trade-offs (T) in 2, and both-worsened (B) in 10 of 42 combinations. Together, these patterns indicate that gender-targeting more frequently narrowed same-axis AUC gaps while modestly widening cross-axis race AUC gaps, whereas TPR gaps showed limited same-axis change with minimal cross-axis disruption.

#### Race-Targeting

In contrast, race-targeting consistently disrupted gender fairness gaps across both metrics. Under AUC (Table 3, Panel B), 2–6 of 7 models showed increased ΔAUC_gender_. Under TPR (Table 4, Race-Targeting, Panel B), 3–4 of 7 models showed increased ΔTPR_gender_ regardless of method. In absolute terms, the largest increases approached 0.050 (EBM under US: 0.004 → 0.053, Δ=+0.050; TabPFN under US: 0.010 → 0.059, Δ=+0.049), with additional sizable increases such as LR under CW (0.008 → 0.052, Δ=+0.044).

Panel C confirmed the asymmetry but revealed metric-dependent dominant patterns. Under AUC, both-worsened was dominant (B=23 of 42): the same interventions failed to narrow even the targeted race AUC gap in the majority of combinations (W=7, T=5, P=7). Under TPR, trade-off was dominant (T=20): same-axis race fairness improved while cross-axis gender fairness worsened (W=16, P=4, B=2). Cross-axis gender harm (T+B) occurred in 28 of 42 combinations under AUC and 22 of 42 under TPR.

The robustness of this asymmetry was supported by 30 random repetitions: all 11 statistically robust gender-targeting cross-axis changes were improvements, while 9 of 12 robust race-targeting cross-axis changes were in the worsening direction.

#### Fairness Metric Divergence

Comparing Panel C across Tables 3 and 4 reveals that cross-axis audit conclusions depend on the fairness metric divergence. Under gender-targeting, the dominant Panel C outcome shifted from trade-off (T=14 under AUC) to paradox (P=18 under TPR), indicating that same-axis AUC gaps were more amenable to improvement than TPR gaps. Under race-targeting, both-worsened was dominant under AUC (B=23) but trade-off under TPR (T=20), indicating that interventions appearing successful under TPR evaluation failed on both axes under AUC. At the individual model level, race-targeting GD in XGBoost showed ΔAUC_gender_ increase (1.3 → 1.4 pp) alongside ΔTPR_gender_ decrease (0.027 → 0.022), an outcome classified as failure under AUC but success under TPR for the same model and method.

### Intersectional Subgroup Analysis

Single-axis balancing methods address disparities along one demographic dimension, but patients exist at the intersection of multiple identities. We examined whether balancing on one axis inadvertently harmed subgroups defined by combinations of gender and race (Figure 2; Tables S8–S9).

**Figure 2.**
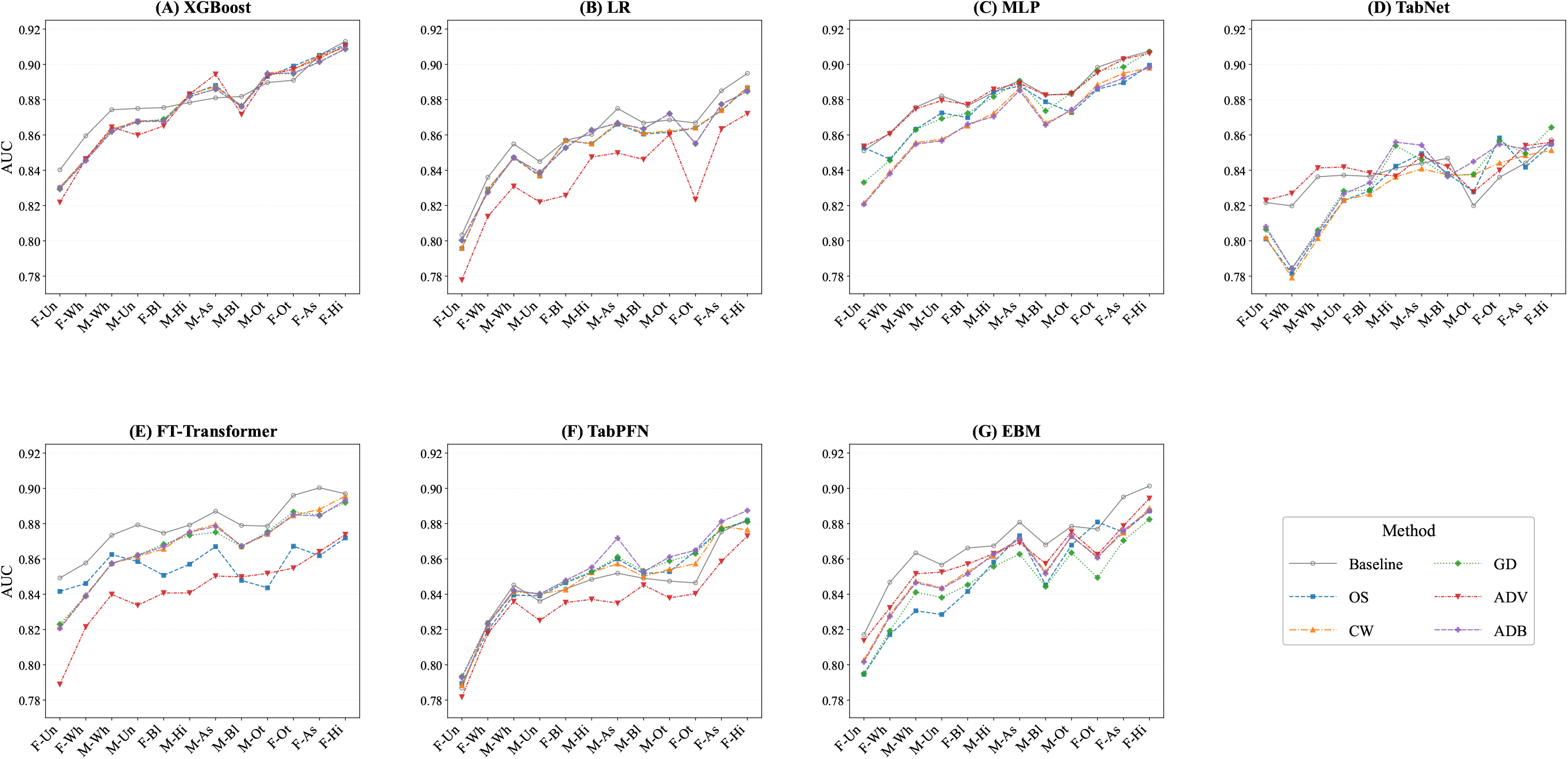
Intersectional subgroup AUC under race-targeting interventions across all seven models. Each panel displays one model; lines represent baseline and five race-balancing methods. Subgroups are ordered by XGBoost baseline AUC (ascending, left = most vulnerable). Native American (*n*=127) and Pacific Islander (*n*=99) subgroups were excluded due to insufficient sample sizes for reliable AUC estimation (*n <* 130; 95% CI width >0.20). Undersampling is excluded due to model collapse in MLP and TabNet (AUC → ~0.50). ADB, Adaptive Balanced Sampling; ADV, Adversarial Debiasing; AUC, Area Under the Receiver Operating Characteristic Curve; BL, Baseline; CW, Class Weighting; F-As, Female Asian; F-Bl, Female Black; F-Hi, Female Hispanic; F-Ot, Female Other; F-Un, Female Unknown; F-Wh, Female White; GD, Group Distributionally Robust Optimization; M-As, Male Asian; M-Bl, Male Black; M-Hi, Male Hispanic; M-Ot, Male Other; M-Un, Male Unknown; M-Wh, Male White; OS, Oversampling. *Alt text: Seven grayscale line charts arranged in a grid, one per model (XGBoost, LR, MLP, TabNet, FT-Transformer, TabPFN, EBM). Each chart plots AUC (y-axis, 0*.*78 to 0*.*92) for 12 intersectional gender-race subgroups (x-axis) under baseline and five race-balancing methods. Subgroups are ordered left to right by ascending baseline AUC: Female Unknown is lowest (approximately 0*.*79–0*.*85) and Female Hispanic is highest (approximately 0*.*86–0*.*91). Six lines per panel represent different methods; they largely follow the same upward trajectory with modest vertical shifts, indicating that balancing methods alter AUC levels but preserve subgroup rank ordering across all models*.

#### Baseline Disparities

Substantial performance variation existed across the 12 intersectional subgroups (2 genders × 6 racial groups) at baseline (Figure 2, baseline lines). AUC gaps (max−min) ranged from 3.7 pp in TabNet (Female Hispanic: 0.857; Female White: 0.820) to 9.6 pp in TabPFN (Female Hispanic: 0.882; Female Unknown: 0.787).

#### Race-Targeting Effects

As shown in Figure 2, race-targeting effects on intersectional disparities varied substantially by model architecture. Race-targeting interventions widened intersectional max−min AUC gaps in 21 of 35 combinations (60%; 7 models × 5 methods, US excluded due to model collapse). TabNet showed gap increases under four of five methods (3.4 to 4.3 pp) and XGBoost under all five (0.6 to 1.6 pp). In contrast, FT-Transformer showed the widest subgroup divergence, particularly under ADV, while TabPFN gaps remained stable despite large baseline disparity (9.6 pp). Even in models where the max−min gap narrowed, Female Hispanic and Female Unknown subgroups predominantly remained the highest- and lowest-performing groups, and the rank ordering of subgroups remained largely unchanged, indicating that single-axis interventions did not redistribute benefits across the full intersectional distribution. Gender-targeting effects on intersectional gaps were minimal (Table S8).

### Calibration

Gender-targeting maintained calibration in most models (mean Brier score change: +0.005), though Tab-Net showed larger increases under advanced methods (up to +0.026 under ADB; Table S7). Race-targeting caused substantially greater calibration loss (mean increase: 0.019). Under race-targeting US, Brier score increased from 0.136 to 0.252 in MLP and from 0.186 to 0.282 in TabNet.

## DISCUSSION

Our results reveal a systematic asymmetry in the cross-axis consequences of demographic balancing (Tables 3 and 4). Gender-targeting interventions largely preserved race fairness, while race-targeting consistently disrupted gender fairness across all six methods and the majority of architectures examined. This asymmetry is invisible to same-axis evaluation alone: each of the 20 trade-off combinations under race-targeting would appear as an un-qualified success if only the targeted axis were assessed, which is the prevailing evaluation practice in clinical ML fairness literature. ^7^ The risk profile of fairness interventions is thus directionally dependent: gender-targeting carries low and largely architecture-specific cross-axis risk, while race-targeting carries systematically higher and more consistent cross-axis risk. This directional asymmetry should be considered a primary evaluation criterion alongside same-axis fairness gains when selecting balancing strategies in clinical settings.

We attribute this asymmetry to differential category structure. Gender comprises two approximately balanced groups (56.7% vs. 43.3%), such that equalizing their representation requires only modest reweighting. Race encompasses eight categories spanning a 438-fold range in sample size, necessitating extreme reweighting that propagates to correlated axes. Gender and race are not statistically independent in this cohort (Figure 1), and the distributional disruption from race reweighting destabilized gender-related model behavior as a byproduct. This propagation was broadly consistent across architectures, from LR to attention-based transformers, suggesting that the effect is not confined to specific model families but reflects a structural property of the balancing task itself. Category complexity, operationalized as the number of groups combined with the degree of imbalance, may therefore serve as a practical predictor of cross-axis risk prior to intervention.

Race-targeting incurred substantially greater performance costs than gender-targeting across both discrimination and calibration. Observed race fairness gap reductions should therefore be interpreted with caution, as gap compression accompanied by overall performance degradation may reflect leveling down; ^27–29^ rather than genuine improvement of disadvantaged groups. This concern is particularly acute under US, where AUC collapsed to near-random levels in two architectures, suggesting that observed TPR gap reductions may partly reflect flattening of predictions rather than genuine improvement of minority-group sensitivity. ^30^ The relative contribution of genuine minority improvement versus overall degradation warrants targeted follow-up.

The same intervention can simultaneously improve fairness on one metric while worsening it on another. Under race-targeting GD, XGBoost showed ΔAUC_gender_ increase (1.3 → 1.4 pp) alongside ΔTPR_gender_ decrease (0.027 → 0.022), an outcome classified as failure under AUC but success under TPR for the same model and method. This divergence is mechanistically grounded: AUC captures ranking performance across all decision thresholds, while TPR is computed at a fixed operating point. These findings reinforce recommendations for decomposed, multi-metric fairness reporting.

Race-targeting widened intersectional max-min gaps in the majority of model-method combinations (Figure 2), and even where gaps narrowed, the rank ordering of subgroups remained largely unchanged, indicating that single-axis interventions did not redistribute benefits across the full intersectional distribution. ^12^ The contrast between XGBoost (consistent gap widening across all five methods) and TabPFN (stable gaps despite large baseline disparity) under identical interventions suggests that architectural inductive biases modulate how distributional perturbations propagate to minority subgroups. These observations support treating intersectional subgroups as an evaluation lens rather than an optimization target; genuinely intersectional improvements likely require intervention designs that simultaneously account for multiple demographic axes during training.

These findings suggest three considerations for fairness-aware clinical AI development. First, cross-axis evaluation may benefit from being integrated as a standard component of pre-deployment fairness auditing. Second, cross-axis evaluation is necessary regardless of method sophistication, as neither simple nor advanced methods consistently avoided cross-axis harm. Third, category complexity along the targeted axis may inform intervention design: when the targeted axis exhibits high cardinality and extreme imbalance, multi-axis methods warrant consideration.

Several limitations should be considered. Our cohort is derived from a single academic medical center (MIMIC-IV), and the observed asymmetry may not generalize to institutions with different demographic compositions. We examined only training-time demographic balancing interventions; post-processing methods may exhibit different cross-axis dynamics. All clinical features were derived from the first 24 hours following ICU admission, and the time from admission to the one-year endpoint varies across patients, potentially introducing differential lead time effects across demographic groups. Additionally, EBM and TabNet were trained on sub-sampled training sets due to computational constraints, which may affect their relative performance estimates. Finally, some intersectional subgroups had extremely small sample sizes, limiting the reliability of subgroup-level fairness estimates for the most vulnerable populations.

## CONCLUSION

This study demonstrates that single-axis fairness interventions produce systematically asymmetric cross-axis effects in clinical risk prediction: gender-targeting largely preserves race fairness, while race-targeting consistently disrupts gender fairness. This asymmetry, attributable to differential category complexity between binary and multi-category demographic axes, is invisible to standard single-axis evaluation and affects the majority of model-method combinations examined. Moreover, observed fairness improvements may reflect leveling down or metric-dependent artifacts rather than genuine equity gains, underscoring the need for multi-metric, cross-axis fairness evaluation as a standard component of pre-deployment auditing in healthcare AI. Future work should develop systematic frameworks for evaluating cross-axis fairness effects across multiple demographic dimensions, and validate these patterns across diverse clinical tasks and institutions.

## Data Availability

The MIMIC-IV (v3.1) database is available from PhysioNet under credentialed access, which requires completion of human subjects research training and a signed data use agreement. Analysis code is available from the authors.

https://physionet.org/content/mimiciv/3.1/

## SUPPORTING INFORMATION

S1 Appendix

### Supplementary methods and results

Outcome selection rationale, metric definitions, hyperparameter configurations, and sensitivity analysis.

**Table S1**. Hyperparameter configurations for all model architectures and balancing strategies.

**Table S2**. Sensitivity analysis: baseline model performance and fairness disparities across three mortality endpoints.

**Table S3**. Overall AUC across all models and balancing methods.

**Table S4**. Subgroup-level AUC across all models and balancing methods.

**Table S5**. Effects of single-axis targeting interventions on same-axis and cross-axis FPR gaps.

**Table S6**. Effects of single-axis targeting interventions on same-axis and cross-axis PPR gaps.

**Table S7**. Subgroup-level Brier score across all models and balancing methods.

**Table S8**. Intersectional subgroup AUC under gender-targeting interventions across all models and balancing methods.

**Table S9**. Intersectional subgroup AUC under race-targeting interventions across all models and balancing methods.

## DATA AVAILABILITY

This study used the MIMIC-IV v3.1 database, available through PhysioNet (https://physionet.org/content/mimiciv/3.1/). ^16^

## AUTHOR CONTRIBUTIONS (CREDIT)

KY: Data curation, Formal analysis, Methodology, Software, Visualization, Writing - original draft. HGK: Conceptualization, Investigation, Methodology, Project administration, Supervision, Validation, Writing - original draft, Writing - review & editing.

## USE OF AI-ASSISTED TOOLS

An AI-based language model (Claude) was used to assist with literature search and manuscript editing (e.g., grammar refinement). Additionally, the model was used to assist with debugging during code development. All outputs were reviewed and verified by the authors.

## REFERENCES

1. Topol EJ. High-performance medicine: the convergence of human and artificial intelligence. Nat Med. 2019;25(1):44–56.

2. Chen IY, Pierson E, Rose S, et al. Ethical machine learning in healthcare. Annu Rev Biomed Data Sci. 2021;4:123–44.

3. Chen RJ, Wang JJ, Williamson DF, et al. Algorithmic fairness in artificial intelligence for medicine and healthcare. Nat Biomed Eng. 2023;7(6):719–42.

4. Ricci Lara MA, Echeveste R, Ferrante E. Addressing fairness in artificial intelligence for medical imaging. Nat Commun. 2022;13(1):4581.

5. Chen F, Wang L, Hong J, et al. Unmasking bias in artificial intelligence: a systematic review of bias detection and mitigation strategies in electronic health record-based models. J Am Med Inform Assoc. 2024;31(5):1172–83.

6. Mehrabi N, Morstatter F, Saxena N, et al. A survey on bias and fairness in machine learning. ACM Comput Surv. 2021;54(6):1–35.

7. Lett E, La Cava WG. Translating intersectionality to fair machine learning in health sciences. Nat Mach Intell. 2023;5(5):476–9.

8. Buolamwini J, Gebru T. Gender shades: Intersectional accuracy disparities in commercial gender classification. In: Conference on Fairness, Accountability and Transparency. PMLR; 2018. p. 77–91.

9. Seyyed-Kalantari L, Zhang H, McDermott MB, et al. Under-diagnosis bias of artificial intelligence algorithms applied to chest radiographs in under-served patient populations. Nat Med. 2021;27(12):2176–82.

10. Kleinberg J, Mullainathan S, Raghavan M. Inherent Trade-Offs in the Fair Determination of Risk Scores. In: Papadimitriou CH, editor. Proceedings of the 8th Innovations in Theoretical Computer Science Conference (ITCS 2017). vol. 67 of Leibniz International Proceedings in Informatics (LIPIcs). Schloss Dagstuhl – Leibniz-Zentrum für Informatik; 2017. p. 43:1-43:23.

11. Colacci M, Huang YQ, Postill G, et al. Sociodemographic bias in clinical machine learning models: a scoping review of algorithmic bias instances and mechanisms. J Clin Epidemiol. 2025;178:111606.

12. La Cava WG, Lee T, Ghassemi M. Intersectional consequences for marginal fairness in prediction models. medRxiv. 2024:2024.01.15.24301365. Preprint.

13. Chen Z, Zhang JM, Sarro F, et al. Fairness improvement with multiple protected attributes: How far are we? In: Proceedings of the IEEE/ACM 46th international conference on software engineering; 2024. p. 1–13.

14. Wang X, Yang CC. Enhancing Multi-Attribute Fairness in Healthcare Predictive Modeling. In: 2025 IEEE 13th International Conference on Healthcare Informatics (ICHI). IEEE; 2025. p. 48–57.

15. Dang VN, Cascarano A, Mulder RH, et al. Fairness and bias correction in machine learning for depression prediction across four study populations. Sci Rep. 2024;14(1):7848.

16. Johnson AEW, Bulgarelli L, Shen L, et al. MIMIC-IV, a freely accessible electronic health record dataset. Sci Data. 2023;10(1):1.

17. Harutyunyan H, Khachatrian H, Kale DC, et al. Multitask learning and benchmarking with clinical time series data. Sci Data. 2019;6(1):96.

18. Zimmerman JE, Kramer AA, McNair DS, et al. Acute Physiology and Chronic Health Evaluation (APACHE) IV: Hospital Mortality Assessment for Today’s Critically Ill Patients. Crit Care Med. 2006;34(5):1297–310.

19. Chen T, Guestrin C. XGBoost: A Scalable Tree Boosting System. In: Proceedings of the 22nd ACM SIGKDD International Conference on Knowledge Discovery and Data Mining; 2016. p. 785–94.

20. Arik SÖ, Pfister T. TabNet: Attentive Interpretable Tabular Learning. In: Proceedings of the AAAI Conference on Artificial Intelligence. vol. 35; 2021. p. 6679–87.

21. Gorishniy Y, Rubachev I, Khrulkov V, et al. Revisiting Deep Learning Models for Tabular Data. Advances in Neural Information Processing Systems. 2021;34:18932–43.

22. Hollmann N, Müller S, Eggensperger K, et al. TabPFN: A Transformer That Solves Small Tabular Classification Problems in a Second. In: The Eleventh International Conference on Learning Representations (ICLR); 2023.

23. Nori H, Jenkins S, Koch P, et al. InterpretML: A Unified Framework for Machine Learning Interpretability. arXiv preprint arXiv:190909223. 2019.

24. Sagawa S, Koh PW, Hashimoto TB, et al. Distributionally Robust Neural Networks for Group Shifts: On the Importance of Regularization for Worst-Case Generalization. In: International Conference on Learning Representations (ICLR); 2020.

25. Zhang BH, Lemoine B, Mitchell M. Mitigating Unwanted Biases with Adversarial Learning. In: Proceedings of the 2018 AAAI/ACM Conference on AI, Ethics, and Society; 2018. p. 335–40.

26. Wastvedt S, Huling JD, Wolfson J. Counterfactual fairness for small subgroups. Biostatistics. 2024.

27. Mittelstadt B, Wachter S, Russell C. The Unfairness of Fair Machine Learning: Leveling Down and Strict Egalitarianism by Default. Michigan Technology Law Review. 2024;30(1). Available from: https://repository.law.umich.edu/mtlr/vol30/iss1/3.

28. Zhang H, Dullerud N, Roth K, et al. Improving the Fairness of Chest X-ray Classifiers. In: Proceedings of the Conference on Health, Inference, and Learning. vol. 174 of Proceedings of Machine Learning Research. PMLR; 2022. p. 204–33. Available from: https://proceedings.mlr.press/v174/zhang22a.html.

29. Pfohl SR, Foryciarz A, Shah NH. An Empirical Characterization of Fair Machine Learning for Clinical Risk Prediction. J Biomed Inform. 2021;113:103621.

30. McCradden MD, Joshi S, Mazwi M, et al. Ethical limitations of algorithmic fairness solutions in health care machine learning. Lancet Digit Health. 2020;2(5):e221–3.

